# Improving the last mile delivery of vaccines through an informed push model: experiences, opportunities and costs based on an implementation study in a rural district in Uganda

**DOI:** 10.1101/2023.11.03.23298045

**Authors:** Pamela Bakkabulindi, Solomon T Wafula, Anthony Ssebagereka, Rogers Sekibira, Aloysius Mutebi, Jimmy Ameny, Christabel Abewe, John Bosco Isunju

## Abstract

**Background:** Many developing countries struggle to attain equitable, timely and efficient availability of potent vaccines at the health facility level. In Uganda, several challenges prevent the reliable distribution of vaccines from the district vaccine store to the health facility level (last mile). The currently practiced mixed push and pull system mode of vaccine delivery is unpredictable, unreliable, and often presents issues of poor vaccine management, vaccine stock-outs and missed opportunities for vaccination.

The overall aim of the study was to improve the efficiency of the last mile delivery of vaccines by implementing an informed push model of vaccine delivery. Specifically, the study aimed to; improve vaccine lead time; standardise cold chain management practices during vaccine transportation; and cost the implementation of the informed push model.

**Methods:** Mixed-methods approach to evaluate the impact of the informed push model on the last mile delivery of vaccines in Gomba district, Uganda was used. Quantitative and qualitative data was collected at baseline and endline. Quantitative data was collected on the mode, frequency, lead time and costs of vaccine delivery; vaccine stock status, and cold chain maintenance of vaccines during transportation using semi-structured interview survey, while the experiences and challenges were explored qualitatively using a guide. Analysis of quantitative data used descriptive statistics and that of costing data used an ingredients approach. Qualitative data using was analysed using a thematic framework.

**Results:** The findings showed that the informed push system improved the efficiency and quality of vaccine delivery at the last mile in Gomba district. The average lead time of vaccine delivery was reduced from 14 days at baseline to 5 days by endline. The number of health facilities reporting timely receipt of vaccines increased from 36.8% at baseline to 100% by endline. Facilities reporting temperature monitoring of vaccines during transit improved from 26.3% at baseline to 100% by endline. Number of health facilities experiencing stock outs reduced from 79% at baseline to 36.8% by endline. The monthly costs incurred by health facilities in vaccine pick up at baseline were $ 170.8. The monthly costs for the informed push model were $445.9 ($ 0.06 per child reached) and more two and half times more than baseline costs of $170.8 incurred by health facilities during pick-up of vaccines from the district vaccine store.

**Conclusion:** The study concluded that informed push model is a financially feasible strategy that could be efficient in improving the vaccine supply chain at the last mile by reducing lead time delivery of vaccines, improving vaccine cold chain management, reducing vaccine stock outs. We recommend the integration of this model into the national immunization program and its subsequent adoption by all districts in Uganda.

## Background

Immunisation is one of the leading cost-effective public health interventions, saving at least two to three million lives each year [1–3]. Sub-Saharan Africa (SSA) faces a disproportionately high burden of vaccine-preventable diseases which cause over 2.4 million deaths among children annually due to limited access to life-saving vaccines. [4].

Many vaccines can now be obtained at low costs and in mass quantities thanks to cutting-edge technology built on many years of research and development and the help of international organisations [2,5]. Over the last decade in developing countries, there have been heightened efforts from UNICEF, Gavi and local governments that have increased vaccine purchases to protect the population from vaccine preventable diseases [6].

Unlike common medicinal drugs, vaccines are highly sensitive to temperature changes and thus need to be maintained at appropriate temperature ranges, outside of which, they lose potency [7].

Globally, the Expanded Programs on Immunization (EPI) rely on well-functioning immunization supply chains systems to ensure that potent vaccines reach the end user in an equitable, timely, and efficient manner [8]. An efficient immunization supply chain remains a critical element in ensuring access to primary health-care services [9]. The ultimate goal and measure of any successful immunization supply chain system is the increased visibility and availability of potent vaccines at the last mile health facility level. This goal still remains hard to attain for many developing countries [10].

Two factors are critical to a functional end-to-end immunization supply chain and logistics systems; a reliable transport system and maintenance of cold chain status of vaccines during storage and transportation [11]. Firstly, a reliable transportation system ensures that vaccines reach the end user in time. Lessons can be learnt from multi-national companies such as Coca-Cola, Johnson & Johnson that have demonstrated their dependability of a reliable transport system for the delivery of goods to the last user [12]. Secondly, a well-maintained cold chain system remains one of the chief cornerstones of a successful vaccination program and not just a high vaccination coverage. Without good cold chain system, a high vaccination coverage is null and without effect [13]. From production to use, vaccines need to be maintained within a certain temperature range [7]. The recommended temperature range are +2°C to + 8°C for refrigerator vaccines, 15°C to - 25°C for freezer vaccines, and −50°C - 80°C for ultra-low temperature vaccines [11].

Like many SSA countries, Uganda struggles to attain the required immunization coverage due to inefficient supply chain systems that often lead to vaccine shortages, missed opportunities for vaccination and resultant vaccine preventable disease outbreaks [6,14]. Indeed, disease outbreaks are largely due to gaps in the herd immunity, which are a consequence of the under-vaccination of the susceptible population [6]. Under-vaccination can result from situations of poor vaccine supply chain systems where vaccines do not reach the target population, stock-out of vaccines or available vaccines that were damaged due to exposure to inappropriate temperatures during transportation [6]. As new vaccines are developed and made available to immunization programs expand, there is a dire and critical need to reduce vaccine wastage, improve efficiencies, reliability, and agility to the immunization supply chain system [15]. Experiences from the recent COVID-19 pandemic have demonstrated a critical need for resilient immunization supply chain systems.

The Ugandan EPI is largely a vertical program that is situated within the Ministry of Health [16] and has the mandate to provide safe, potent, and effective vaccines for all children and women of childbearing age so as to reduce morbidity, mortality, and disability due to vaccine-preventable diseases [17]. Since 2012, the key functions of vaccine forecasting, and procurement became shared responsibilities between the EPI and National Medical Stores (NMS). However, other roles including storage at the national level and distribution to all district vaccine stores countrywide were given to NMS to capitalize on their larger storage and transportation infrastructure. At the district level, the district cold chain technicians (DCCT) assume responsibility for cold chain equipment maintenance as the primary role though they also support proper vaccine storage and handling and in some cases distribution of vaccines directly to the health facilities.

Although Uganda has made considerable improvements in her vaccine supply chain, there is still room to improve her vaccine delivery at the last mile (from the district vaccine store to health facility level). Several bottlenecks are reported to prevent the reliable distribution of vaccines and immunization supplies at the last mile [16]. In the current Uganda vaccine delivery model (Fig1), the National Medical Stores executes the delivery of vaccines up to the district vaccine store (DVS). From the DVS to health facilities is a mix of push and pull system, with the pull system (health facilities picking their stock) being the dominant one where approximately 85% of the health facilities collecting vaccines from the DVS [16] which ad hoc collection system often results in inconsistent availability of vaccines at the DVS and the health facility level [16].

**Fig 1:**
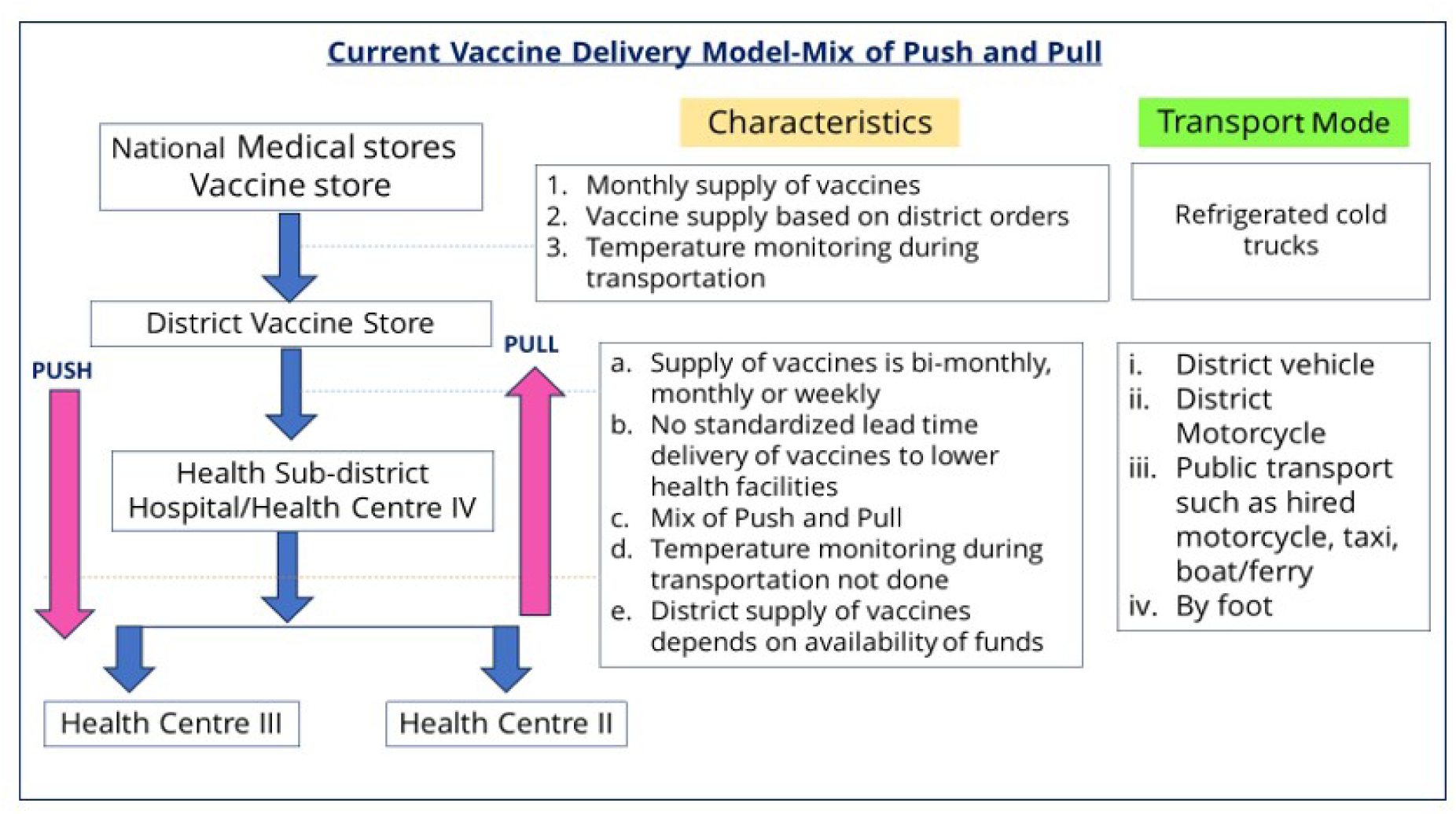
Current vaccine delivery model.

The currently practiced mixed system of push and pull vaccine delivery model is unpredictable, unreliable and often presents issues of poor vaccine management, vaccine stock-outs and missed opportunities for vaccination at the health facilities. The district has no visible role on the equitable distribution of vaccines to each health facility. Therefore, the district is unable to accurately forecast and quantify its vaccine needs and only must be dependent on the health facility forecasts. Also, the supervision of vaccine management of vaccine management by the DCCT at the health facilities is hardly done [16]. A key financing gap cited at the district level is the lack of funds to transport vaccines from the district vaccine store to health facilities [17].

There is paucity in information on the intricacies, opportunities and costs involved in the last mile delivery of vaccines in Uganda. There is a need to establish the number of days required to distribute vaccines, the experiences of cold chain maintenance during transportation of vaccines and the costs involved in the last mile vaccine delivery of vaccines. The overall aim of the study was to improve the efficiency of the last mile delivery of vaccines by implementing an informed push model of vaccine delivery. Specifically, the study aimed to; improve vaccine lead time; standardise cold chain management practices during vaccine transportation; and cost the implementation of the informed push model.

## Methods

### Study setting

The study was implemented in a rural district called Gomba, located in the central region of Uganda, 95km from the capital district, Kampala. The study was conducted between February and August 2022. As of 2020, Gomba district had a population of 169,518 people; 34,243 children under-five years; 19 health facilities offering vaccination services; Measles Containing Vaccine (MCV) coverage of 55%; Bacillus Calmette-Guerin (BCG) vaccine coverage of 65%; and had experienced 2 measles outbreaks in that same year.

Uganda’s health care system has four levels: hospitals and health centres II, III, and IV. Hospitals are higher-level facilities with consultant physicians and specialized services. Health centres II, III, and IV are lower-level facilities (or primary health care) that provide basic services at parish, sub-county, and county levels, respectively. For instance, health centre IIs offer outpatients consultations, health centre IIIs offer some inpatient care and some laboratory services, and health centre IVs offer caesarean deliveries and blood transfusion services [18]. However, all these facilities offer immunization services.

### Study design

The study consisted of pre and post cross-sectional study design that employed quantitative and qualitative methods of data collection. After collection of data at baseline, we implemented an informed push model of vaccine delivery in all the health facilities (19) offering vaccination services in Gomba district.

### Description of the informed push model intervention

The informed push model (IPM) intervention was implemented in all the 19 government health facilities providing immunisation services in Gomba district from March to May 2022. With this intervention, the vaccines were delivered monthly from the District Vaccine Store (DVS) to all the 19 immunizing health facilities (Fig 2).

**Fig 2:**
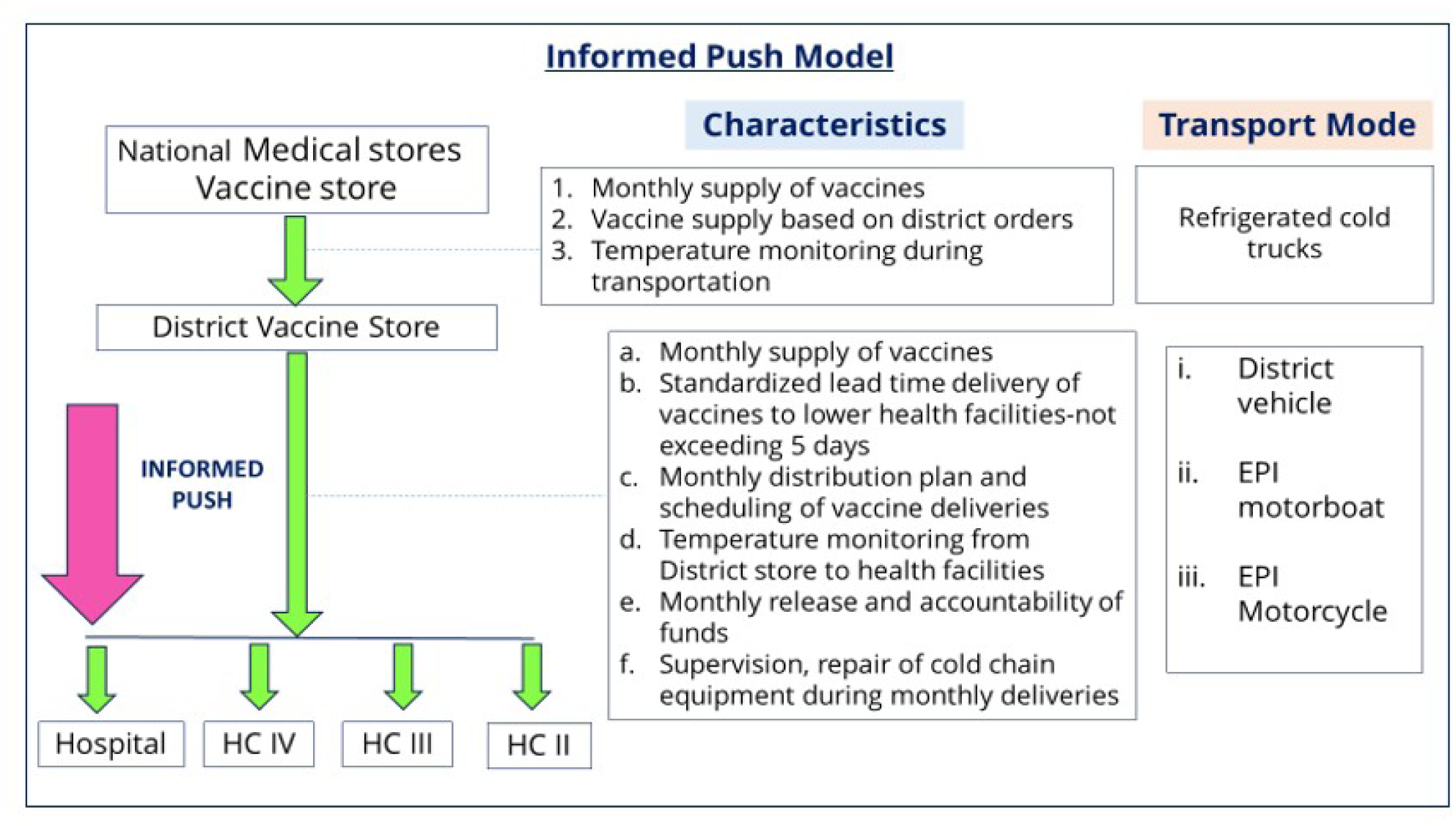
Informed Push Model.

The IPM model differs from an ordinary push model in that it includes an element of first obtaining information on vaccine stock status from the EPI health worker before delivery of the vaccines. During the implementation of the IPM, the DCCT conducted the following tasks: 1) Contacting all the 19 health facilities to determine and confirm the status of their vaccine stock; 2) Designing a distribution plan and strategy of vaccine delivery and thereafter moving according to this planned schedule to each health facility; 3) Maintaining the cold chain of vaccines by use of conditioned icepacks; 4) Monitoring and record the temperature of vaccines at specified intervals (at start of journey and at the point of delivery at a health facility). In this study, during transportation of vaccines, the acceptable temperature range was considered +2°C to + 8°C and any temperatures outside this range had to be flagged to the study investigators; 5) Recording of the routes used, fuel consumed, and kilometres travelled; 6) Documenting the lead time delivery of vaccines which is described as the number of days taken to deliver vaccines to all health facilities in each month.

For the intervention period, funds were given monthly for a period of 3 months to cover costs for 1) fuel for transportation of vaccines; 2) safari day allowances for the vaccine delivery team of 3 people who were DCCT, District cold chain assistant (DCCA), and driver; 3) car maintenance costs and, 4) airtime for communication and coordination with the health facility teams. There was no need for vehicle hire as there was an existent district owned vehicle tagged for immunization activities.

### Data collection

Experienced research assistants were trained for 5 days to administer the quantitative survey questionnaire and qualitative guide. Quantitative and qualitative data was collected before (February 2022) and after (August 2022) the implementation of the study interventions. The study targeted a total of 20 respondents who included one DCCT based at the district level and 19 EPI health workers based at the health facility level for both the survey and qualitative interviews.

Quantitative survey data was collected using a pre-tested structured questionnaire. Questions included sociodemographic characteristics; vaccine management training; mode of vaccine delivery; vaccine delivery schedules; lead time delivery of vaccines; vaccine ordering; vaccine stock-outs; functional cold chain equipment; cold chain management during vaccine transportation; and costs involved in vaccine delivery/pick up.

The qualitative guide included questions on the processes and experiences on; vaccine delivery/pickup; management of cold chain during vaccine delivery/pickup; vaccine stock-outs; costs involved in vaccine pick-up/delivery; challenges and opportunities for last mile vaccine delivery.

### Outcomes measures

The primary outcomes for the study were lead time delivery of vaccines; the costs incurred to deliver vaccines to every health facility and maintenance of vaccine cold chain temperature.

The secondary outcomes included frequency of vaccine stock-outs; functionality of health facility cold chain equipment; the preferred mode of vaccine delivery; the number of of health workers mentored on vaccine forecasting and management.

### Data management and analysis

#### Quantitative data

Quantitative data was collected using the Kobo tool (Kobo Inc., Boston, MA, USA) and exported to Excel. Then data was reviewed for completeness, coded, and later analysed using Stata 15 (Stata Crop., College Station, TX, USA). Frequencies, means, and standard deviation were used to summarize the data. The analysis was stratified by pre (February 2022) and post intervention (August 2022). Due to the small sample size of respondents, comparisons were based on change in frequencies.

The analysis of the costing data used an incremental ingredients approach [19,20] where each resource input was identified, quantified, and valued by multiplying the quantity of inputs by their unit prices. A provider’s perspective was considered, and the cost analysis is presented by two cost centres: the costs incurred by health facilities as they pick vaccines and (b) the costs incurred by the district vaccine store for delivery of vaccines.

Given that an incremental costing was conducted, the study did not consider already sunken indirect health system costs such as human resources, buildings, purchase of capital equipment, volunteer time, and other opportunity costs. Therefore, the resource inputs considered for the incremental cost analysis were those additional resources that would be required to facilitate the push model for vaccine supply chaining including transport, fuel, vehicle hire, repairs and maintenance costs, and communication costs.

Cost data was captured in Uganda shillings and later expressed in US Dollars (USD). The exchange rate at the time of the study was 1 USD being equivalent to 3,700 Uganda shillings as of year 2022. Sensitivity analyses were carried out to assess the extent to which uncertainty in some variables affected the study results.

#### Qualitative data

For qualitative data, audio tape recordings were all together transcribed verbatim into English by the same research assistants who conducted the interviews, while preserving the local language concepts. In keeping with this methodology, the transcripts were read and re-read to obtain a good understanding of the data. A thematic framework was identifying by writing memos in form of short phrases, ideas or concepts arising from the data in the margins of the text [21]. These were then organized according to specific categories. This was followed by highlighting and sorting out quotes and making comparisons within and between cases. The quotes were then lifted from their original context and re-arranged under the newly developed themes [21]. Finally, the data was interpreted based on internal consistency, frequency and extensiveness of responses, specificity of responses and trends or concepts that cut across the various discussions. The analyzed data was then presented in text form.

#### Ethical considerations

The study was approved by Makerere University School of Public Health Higher degrees, Research and Ethics committee (SPH-2021-138: A) and the Uganda National Council of Science and Technology (HS2183ES). Permission to carry out the study was received from Gomba district leaders. Written consent was obtained from each respondent.

## Results

The study intervention was conducted for all nineteen health facilities offering immunisation services in Gomba district. As seen in table 1, the majority of the respondents were EPI focal persons (42.1%) who were enrolled nurses by profession. Most of the respondents (73.7%) had been trained on vaccine management in the past five years. The health facilities visited were mostly of level III (47.4%) and level II (47.4%). Many of the health facilities (26.3%) were within the 21-30 kilometre and notably two health facilities (10.5%) were within the farthest distance of 71-80 kilometres. Most of the health facilities (89.5%) were government owned (table 1).

**Table 1:** Sociodemographic characteristics of respondents in Gomba district, Uganda.

| Characteristics | Number of Participants<br>N = 19 | Percentage |
| --- | --- | --- |
| ***Designation of respondents*** |  |  |
| EPI Focal Person | 8 | 42.1 |
| Health Facility In-Charge | 6 | 31.6 |
| Others*<sup>1</sup> | 5 | 26.3 |
| ***Profession of respondents*** |  |  |
| Enrolled Nurse | 8 | 42.1 |
| Midwife | 4 | 21.1 |
| Nursing assistant | 3 | 15.8 |
| Registered Nurse | 2 | 10.5 |
| Other*<sup>2</sup> | 2 | 10.5 |
| ***Ever received any training on vaccine management practices in the past 5 years*** |  |  |
| Yes | 14 | 73.7 |
| No | 5 | 26.3 |
| ***Years in service provision of health workers, mean (SD)*** |  |  |
| In general health service | 10.74 (6.67) |  |
| In immunization service | 9.37 (6.94) |  |
| ***Type of Health Facility*** |  |  |
| Health Centre II | 9 | 47.4 |
| Health Centre III | 9 | 47.4 |
| Health Centre IV | 1 | 5.3 |
| ***Health facility ownership*** |  |  |
| Government | 17 | 89.5 |
| Private | 2 | 10.5 |
| ***Distance of the health facilities from the district*** |  |  |
| 0-10 kilometers | 1 | 5.3 |
| 11-20 kilometers | 3 | 15.8 |
| 21-30 kilometers | 5 | 26.3 |
| 31-40 kilometers | 4 | 21.1 |
| 51-60 kilometers | 2 | 10.5 |
| 61-70 kilometers | 2 | 10.5 |
| 71-80 kilometers | 2 | 10.5 |
| *<sup>1</sup>Others included Health assistant, enrolled nurse, enrolled midwife, vaccinator and laboratory assistant |  |  |
| *<sup>2</sup>Others included Health assistant and laboratory assistant |  |  |

### Mode and frequency of vaccine delivery

As seen in table 2, at baseline, a mix of push and pull mode of vaccine delivery existed with pull mode being the dominant one (79%) with only 7/19 (36.8%) facilities reporting a push model. However, the IPM model was implemented during the intervention and all 19 facilities (100%) reported a push model at endline.

**Table 2.**
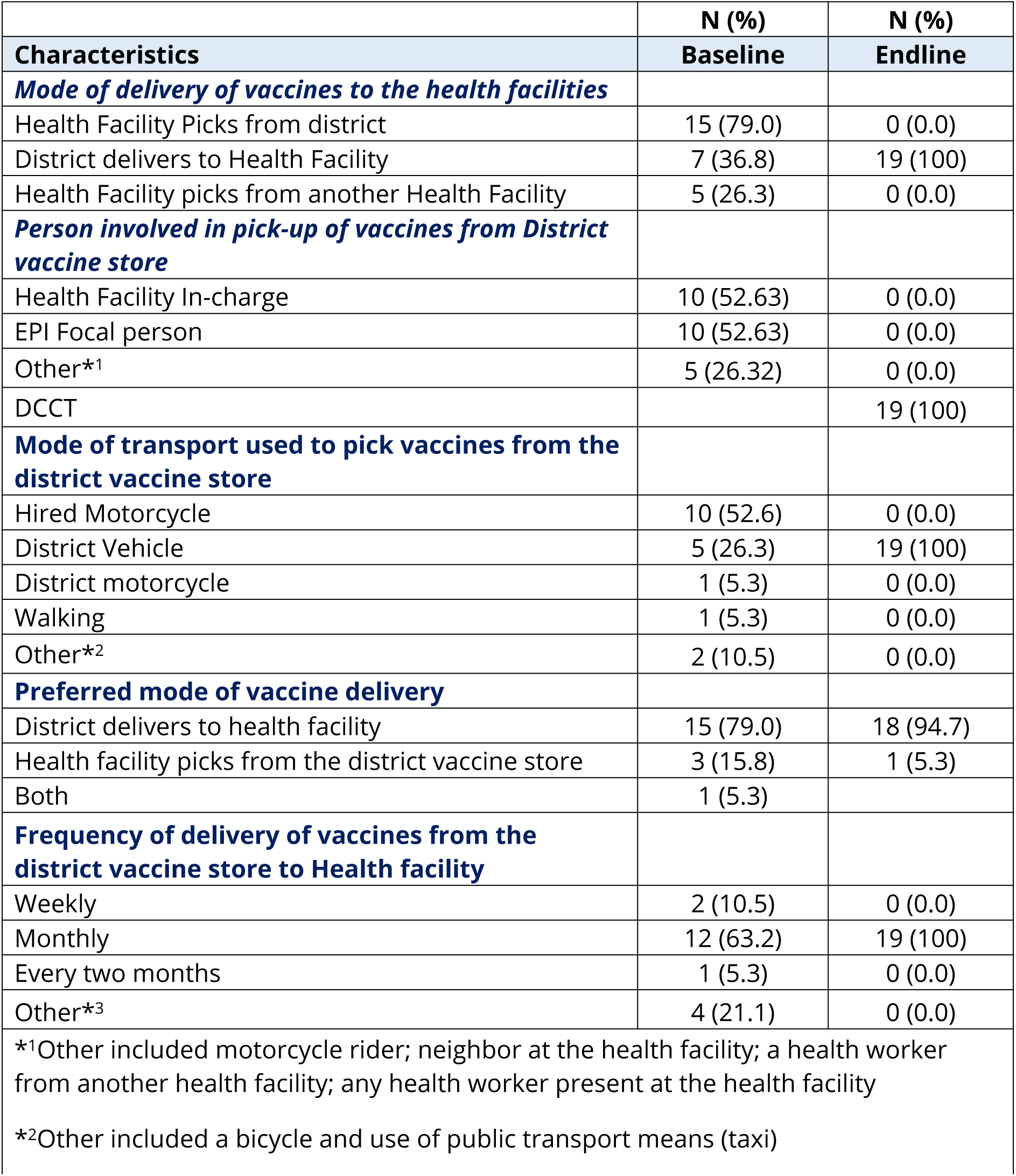
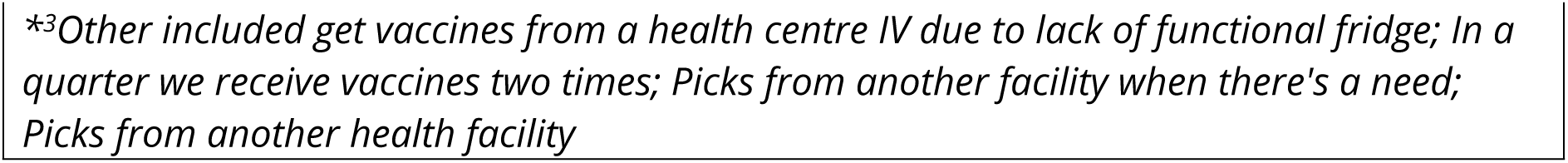
Mode and frequency of delivery of vaccines in Gomba district, Uganda.

At baseline, the health facility in-charges (52.63%) and/or EPI focal persons (52.63%) were involved in vaccine pick-up. However, we note unqualified persons (26.3%) picking vaccines such as motorcycle riders. By endline, the DCCT was the one delivering vaccines to all the 19 immunizing health facilities as per the IPM (table 2).

The reported commonest mode of transport used to pick vaccines from the district vaccine store at baseline were hired motorcycles (52.6%) as compared to the district vehicle (100%) at endline (table 2).

At baseline only 12 (63.2%) facilities reported receiving vaccines at a monthly frequency, and this changed to all the 19 (100%) facilities being supplied monthly with IPM.

Majority of health workers at baseline and endline (79.0% and 94.7% respectively) preferred the district to deliver vaccines (table 2).

### Finances and stock out challenges influence the dynamics involved in delivery dynamics

Qualitative interviews also pointed out some difficulties with modes of vaccine delivery and how this was due to limited finances and consequently caused understocking or sometimes overstocking as remarked by the district cold chain technician and one EPI focal person:

> “Before implementation of the study, most health facilities picked vaccines from the district vaccine store at their convenience. This was mainly because there were no available financial resources to be used for delivery of vaccines.” When health facilities pick vaccines, there is tendency for them to take more than they need which leads to overstocking on their part and understocking for the health facilities that pick last.” **KII, District Cold Chain Technician, Gomba district.**

> “The reason why we pick vaccines from the district store is because the district does not deliver them to the facility, so instead of staying without vaccines, one must spend money on transportation to ensure that vaccines are obtained”. **KII-12, Health worker, Gomba district.**

### Informed push delivery a one stop solution to various challenges at the health facility level

Respondents also thought IPM may solved challenges of understocking in some facilities and overstocking in other facilities as it is common with earlier mixed push and pull system.

> “From my experience, it is better for the cold chain technician to distribute vaccines because then it allows for efficient distribution as I would first quantify and confirm the vaccine needs of a health facility before distribution. I was even able to re-distribute vaccines between facilities which prevented over/under stocking. I was also able to identify cold chain challenges and solve those I could there and then at the health facility.” **KII, District Cold Chain Technician, Gomba district.**

> “The best mode of vaccine delivery is when the district delivers to the health facility. This is because most of the time it is cost effective because the distance from the health center to the district is a long distance that means you have to use much transport and you again have to facilitate the person who is going to pick the vaccine so it is less expensive when the district delivers, it would cost less when the district supplies all facilities rather than everyone going there to pick”. **KII-4, Health worker, Gomba district.**

Health workers complained of the inconveniences that are associated with vaccine pick-up which sometimes results in not being able to attend to patients / clients. One respondent remarked as follows

> “Picking of vaccines is inconveniencing for the health workers and patients. For example, a health facility at the level of health center II has three staff so when one leaves then workload increases for the two who have remained at the facility”. **KII-9, Health worker, Gomba district.**

> “Vaccine pick up is difficult for us. First of all, our health facility is in a remote location, and it is far from where you find a taxi costs three dollars and again the transport to here is in a taxi is not readily available and is only in the evening, so you end up walking that distance at night”. **KII-5, Health worker, Gomba district.**

> “When the district brings, we are sure that the vaccines will be received in good condition. We also save on transport and reduce on the waiting time for mothers because then the health worker does not have her duties to pick vaccines”. **KII-2, Health worker, Gomba.**

### Vaccine delivery lead-time

When asked the actual number of days in each month recommended for vaccine delivery also known as the lead time, they reported incorrectly 22 days at baseline and correctly within 5 days at endline (table 3).

> Qualitative findings also corroborated this “Before implementation of the study, it would take an entire month for the health facilities to pick vaccines from the district vaccine store as and when they had the time or felt it convenient”. Given that funds were readily available during the implementation, we took 5 days to distribute vaccines to all health facilities.” **KII, District Cold Chain Technician, Gomba district.**

**Table 3:**
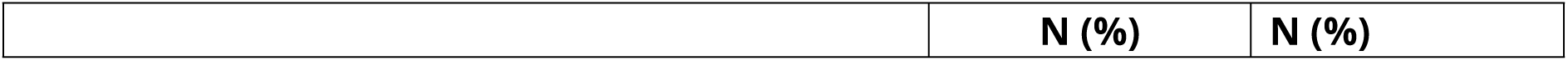
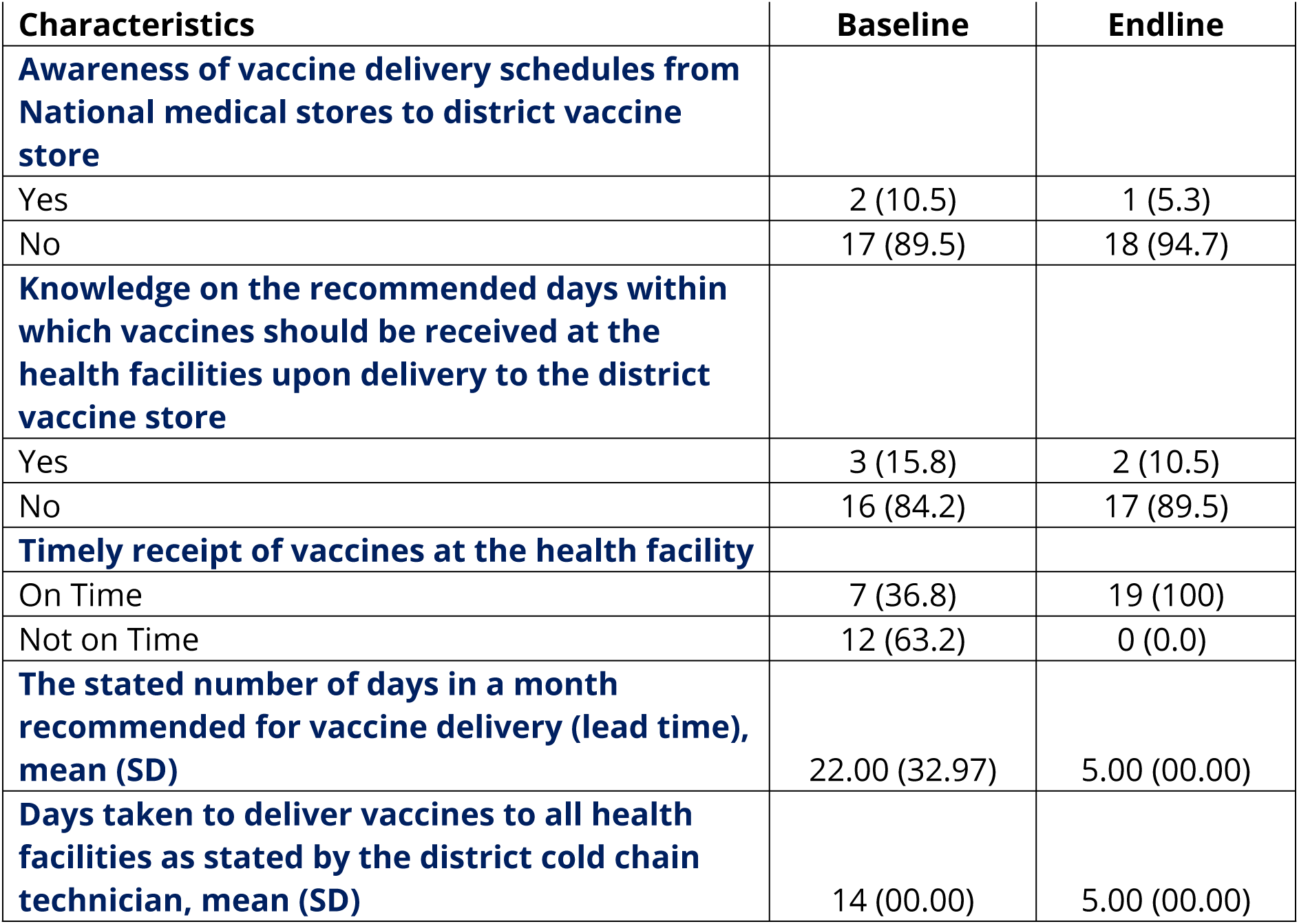
Lead time delivery of vaccines in Gomba district, Uganda.

| Characteristics | Baseline | Endline |
| --- | --- | --- |
| **Awareness of vaccine delivery schedules from National medical stores to district vaccine store** |  |  |
| Yes | 2 (10.5) | 1 (5.3) |
| No | 17 (89.5) | 18 (94.7) |
| **Knowledge on the recommended days within which vaccines should be received at the health facilities upon delivery to the district vaccine store** |  |  |
| Yes | 3 (15.8) | 2 (10.5) |
| No | 16 (84.2) | 17 (89.5) |
| **Timely receipt of vaccines at the health facility** |  |  |
| On Time | 7 (36.8) | 19 (100) |
| Not on Time | 12 (63.2) | 0 (0.0) |
| **The stated number of days in a month recommended for vaccine delivery (lead time), mean (SD)** | 22.00 (32.97) | 5.00 (00.00) |
| **Days taken to deliver vaccines to all health facilities as stated by the district cold chain technician, mean (SD)** | 14 (00.00) | 5.00 (00.00) |

The lead time delivery of vaccines improved from 14 days at baseline to 5 days at endline. Similarly, timely receipt of vaccines at the health facilities improved from 36.8% at baseline to 100% by endline (table 3).

Health worker knowledge of the correct days for lead time delivery of vaccines improved from incorrect reported 22 days at baseline to correct 5 days at endline (table 3).

As seen in table 3, by baseline and endline, most of the respondents (89.5% and 94.7% respectively) were still not aware of the vaccine delivery schedules from the national stores to the district vaccines stores.

This information is collaborated by the qualitative interviews:

> “No, we are not aware of the vaccine delivery times, though sometimes we are informed but at times we just go there not knowing the vaccine delivery schedule for national medical stores”. **KII-1, Health worker, Gomba.**

> “The challenge is we are not informed about the delivery of vaccines by national medical stores because if we knew then we would go there at the right time, but unfortunately, we always go to the district vaccine store when the vaccines are already finished”. **KII-7, Health worker, Gomba.**

### Cold chain and Vaccine management

Notably at baseline, only 5 (26.3%) of respondents reported that they were monitoring temperature of vaccines during transportation. However, during implementation of the study, the DCCT was mandated to monitor temperature twice (at the start and end of the journey) of the vaccines during vaccine transportation. This is reflected in the endline results as all the respondents (100%) attested that temperature monitoring for vaccines was conducted during their transportation (table 4).

**Table 4:** Cold Chain and Vaccine Management processes in Gomba district, Uganda.

|  | N (%) | N (%) |
| --- | --- | --- |
| Characteristics | Baseline | Endline |
| **Availability of cold chain storage equipment at health facility** |  |  |
| Yes | 17 (89.5) | 19 (100) |
| No | 2 (10.5) | 0.0 (0) |
| **Functionality of the available cold chain equipment** |  |  |
| Yes | 14 (73.7) | 19 (100) |
| No | 3 (15.8) | 0.0 (0) |
| **Type of available functional cold Chain equipment** |  |  |
| Fridge | 17 (89.5) | 19 (100) |
| Vaccine carriers | 8 (42.1) | 14 (73.4) |
| Fridge tag | 5 (26.3) | 11 (57.9) |
| Cold boxes | 2 (10.5) | 4 (21.1) |
| Freezers | 1 (5.3) | 4 (21.1) |
| Other*1 | 1 (5.3) | 2 (10.5) |
| **Type of cold chain equipment used for storage of vaccines during transportation** |  |  |
| Vaccine Carrier | 19 (100) | 0 (0.0) |
| Cold box | 0 (0.0) | 19 (100) |
| **Type of icepacks used during vaccine transportation** |  |  |
| Frozen icepacks | 13 (68.4) | 0 (0.0) |
| Conditioned icepacks | 6 (31.6) | 19 (100) |
| **Temperature monitoring during vaccine transportation from the district vaccine to health facilities** |  |  |
| Yes | 5 (26.3) | 19 (100) |
| No | 14 (73.7) |  |
| **Type of temperature monitoring tool used during vaccine transportation for those that carried out monitoring** |  |  |
| Thermometer | 3 (15.8) | 0 (0.0) |
| Fridge tag | 2 (10.5) | 19 (100) |
| **Frequency of monitoring temperature during transportation of vaccines from district vaccine store** |  |  |
| Not monitored | 11 (57.9) | 0 (0.0) |
| Once | 3 (15.8) | 0 (0.0) |
| Twice | 2 (10.5) | 19 (100) |
| Do not know | 3 (15.8) | 0 (0.0) |
| **Issues with vaccines received** |  |  |
| Yes | 3 (15.8) | 2 (10.5) |
| No | 16 (84.2) | 17 (89.5) |
| **Type of issues with vaccines received** |  |  |
| Vaccines with vaccine vial monitor changes | 2 (10.5) | 1 (5.3) |
| Poor bundling | 1 (5.2) | 1 (5.3) |
| **Health worker knowledge of reading and interpreting VVM changes** |  |  |
| Yes | 19 (100) | 19 (100) |
| **Health worker checking for VVM changes at the point of receipt of vaccines** |  |  |
| Yes | 18 (94.7) | 16 (84.2) (16) |
| No | 1 (5.3) | 3 (15.8) |

\*<sup>1</sup> Other included icepacks

More people indicated, they didn’t monitor temperature prior to implementation of IPM:

> “To be honest, I do not monitor for temperature when I go to pick vaccines from the district vaccine store.” **KII-17, Health worker, Gomba.**

> “I won’t lie to you that we do it. Because we use a taxi or motorcycle to pick vaccines from the district vaccine store to the health facility. So, with that mode of transport, you can’t monitor the temperature.’’**KII-2, Health worker, Gomba.**

> “To be honest, before implementation, during the vaccine delivery, I would measure the temperatures but not record them anywhere. But during implementation, I was mandated to record the temperatures before, during and on arrival at the health facilities, this was big lesson for me.” **KII, District Cold Chain Technician, Gomba district.**

The availability of cold chain equipment for storage of vaccines improved from 89.5% at baseline to 100% at endline (table 4). Similarly, the functionality of this equipment improved from 73.7% at baseline to 100% at endline. This is because the DCCT was able to repair broken down fridges during the monthly delivery of the vaccines (table 4).

At baseline, most of the respondents (68.5%) incorrectly reported using frozen icepacks (placed within vaccine carriers) during vaccine transportation. With the DCCT being able to observe the right practices, this was corrected and by endline, all the respondents (100%) reported that use of conditioned icepacks during vaccine transportation (table 4).

The number of respondents reporting cold chain issues with vaccines received decreased from 15.8% (3/19) at baseline to 10.5% (2/19). The commonest issue reported was changes in the vaccine vial monitor (VVM). In addition, all the health workers knew how to read and interpret the vaccine vial monitor (VVM) changes and more than 80% of the respondents checked for VVM changes upon receipt of vaccines at baseline and endline (table 4).

Key informants highlighted some advantages of the IPM approach. One respondent remarked as follows:

> “The push system or when the district delivers vaccine had so many advantages. We were able to identify faulty fridges and have them repaired in a short time. We were able to distribute vaccines according to the rightful need. We were able to train the health workers in quantification, cold chain maintenance and record keeping. At some point, we even re-distributed vaccines to ensure a good stock balance. We noticed an improvement in the number of children immunized during the 3 months of implementation.’ **KII, District Cold Chain Technician, Gomba district.**

When asked about the earlier strategy for vaccine delivery at baseline, respondents highlighted the difficulty in maintenance of the cold chain during transit and yet this had detrimental practice could affect the vaccine potency. One respondent stated as follows:

> “…., in my opinion, there is no proper cold chain maintenance when we pick vaccines from the district store. One can go with icepacks in the vaccine carriers. But by the time we get at the district store, they have melted with hardly any cold temperature. So, am not sure that col chain is maintained as we transport these vaccines” **KII-5, Health worker, Gomba.**

### Vaccine stock status

All the respondents at both baseline and endline reported the availability of the issuing and ordering tool (table 5). The number of health facilities reporting that they did not receive vaccines as ordered reduced from 57.9% at baseline to 26.3% at endline. Similarly, the number of health facilities experiencing vaccine stock outs also reduced from 79% at baseline to 36.8% at endline following implementation of IPM.

**Table 5:** Vaccine Stock status, Gomba district, Uganda.

|  | **N (%)** | **N (%)** |
| --- | --- | --- |
| **Characteristics** | **Baseline** | **Endline** |
| **Availability of issuing and ordering tool at health facility** |  |  |
| Yes | 19 (100) | 19 (100) |
| **Health worker Knowledge on use of the issuing and ordering tool** |  |  |
| Yes | 19 (100) | 19 (100) |
| **Health Facility receiving vaccines as ordered** |  |  |
| Yes | 8 (42.1) | 14 (73.7) |
| No | 11 (57.9) | 5 (26.3) |

| **Health facility experiencing stock out of vaccines** |  |  |
| --- | --- | --- |
| Yes | 15<br>(79.0) | 7 (36.8) |
| No | 4 (21.1) | 12<br>(63.2) |
| **Instances of under-fulfilled orders received by health facility** |  |  |
| Yes | 12<br>(63.2) | 6 (31.6) |
| No | 7 (36.8) | 13<br>(68.4) |

As seen in table 5, all the respondents at both baseline and endline reported the availability of the issuing and ordering tool at the health facility. However, at baseline, more than half of respondents (57.9%) reported that their health facilities did not receive vaccines as ordered which picture improved at endline as only 26.3% reported not receiving vaccines as ordered.

Similar disparities between number of vaccines ordered and supplied was highlighted qualitatively at baseline as indicated as follows:

> “We usually do not receive the same quantity of vaccines as ordered for. Sometimes I reach district vaccine store and find that some vaccines are out of stock, and I end up taking less quantities than what is needed”. **KII-8, Health worker, Gomba**

> “Other times on arrival at the district vaccine store, I find most vaccines are taken and are finished so I take what is left.” **KII-13, Health worker, Gomba**

### Costs incurred during vaccine delivery

#### Summary of Cost Analysis Results

The costs incurred by the health facilities as they picked vaccines from the DVS at baseline were $170.8 as compared to those incurred by the district as they delivered vaccines to health facilities which were $324. At endline, the health facilities did not incur any costs while the district incurred $ 445.9 (table 6).

**Table 6:** Summary of costs incurred by health facilities and the district in vaccine delivery.

| Resource Input | Costs incurred by Health Facilities in USD |  | Costs incurred by District in USD |  |
| --- | --- | --- | --- | --- |
|  | Baseline | Endline | Baseline | Endline |
| Transport/Fuel | 170.8 | 0 | 324 | 270 |
| Repairs & maintenance | 0 | 0 | 0 | 81 |
| Safari Day Allowance | 0 | 0 | 0 | 81 |
| Communication | 0 | 0 | 0 | 14 |
| **Total** | **170.8** | **0** | **324** | **445.9** |

### Baseline costs incurred by health facilities for Vaccine Pick-up

At baseline, the combined costs incurred for pick-up of vaccines from the DVS for all the 19 health facilities were $170.81. $ 21.62 was the highest amount paid by a health facility and the lowest being $3.24 (table 7).

**Table 7:**
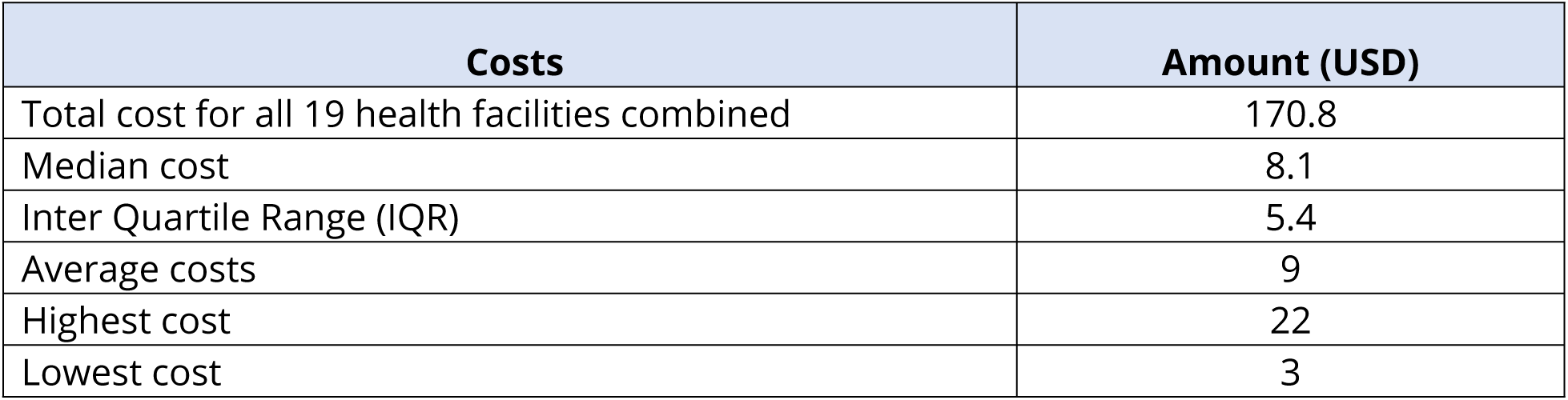
Baseline monthly average costs incurred by health facilities for vaccine pick-up.

The major areas of expenditure were transport (53%) and or fuel (42%). In addition, a significant 32% under ‘other cost’ included cost areas for hiring of motorcycles, motorcycle repairs, lunch, and airtime for communication. The respondents indicated that the primary health care funds (89%) were the major source of funds for vaccine pick-up. However, 21% of the respondents reported using out-of-pocket funds for vaccine pick-up from multiple choice questions asked (Fig 3).

**Fig 3:**
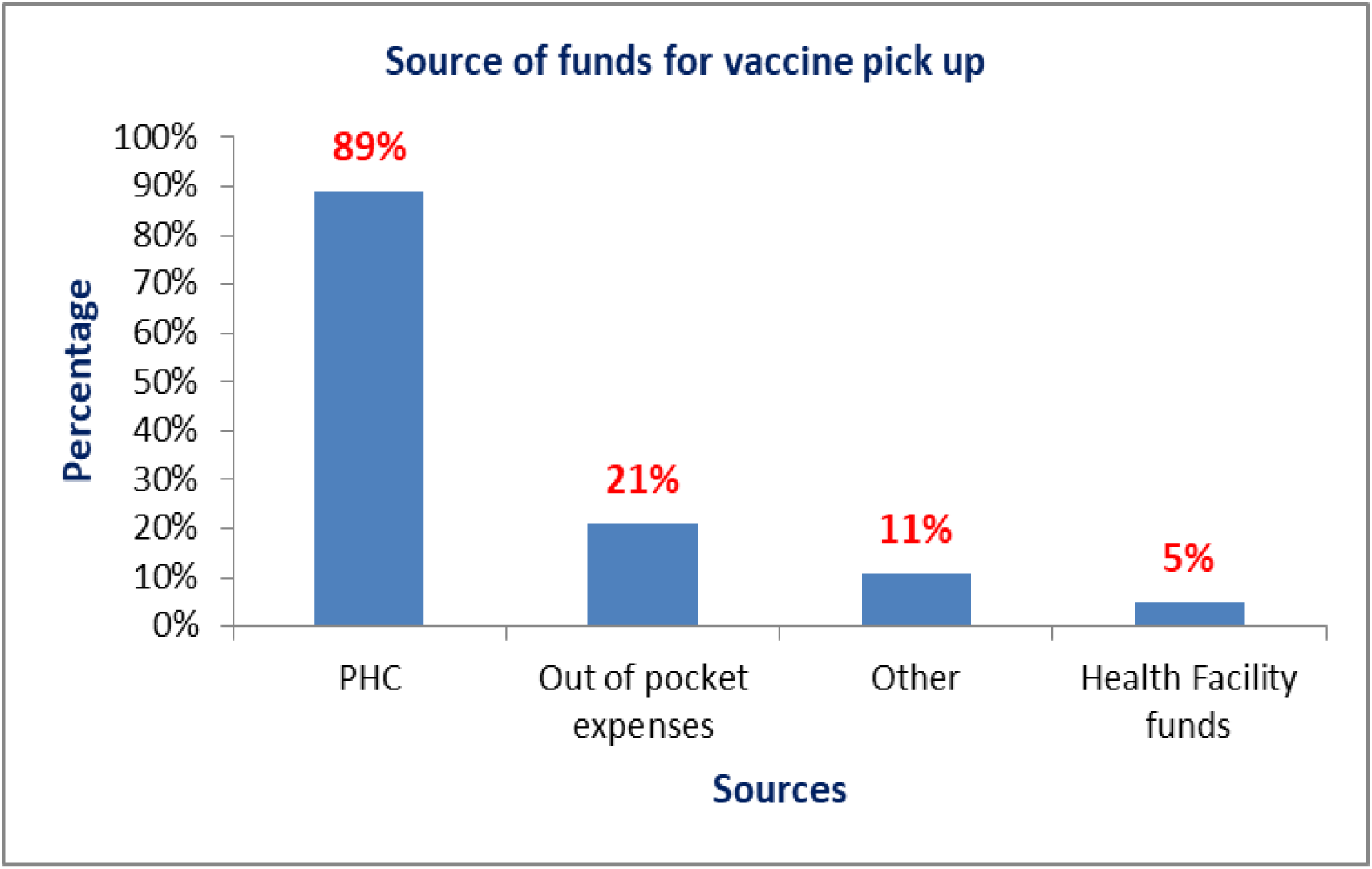
Graph showing source of funds for vaccine pick-up at baseline.

Qualitative interviews also provided perspective on source of funds as seen *below:*

> “The source of funds for transport to pick vaccines is a charge we put to the clients. We get the money from mothers who come for immunization. Mothers are charged $ 0.3 each time they come for immunization so it’s the money we use for transport costs during vaccine pick-up”. **KII-6, Health worker, Gomba.**

### Costs incurred by district to deliver vaccines to all health facilities during implementation of IPM

Prior to the study implementation, there were no consistent district budgeted costs for vaccine delivery. However, the average baseline costs were $ 324 and these catered for only fuel costs for delivery of vaccines. The push model required the district cold chain team to deliver vaccines to all the 19 immunising health facilities using the district car that was allocated to the EPI department. There were three personnel involved in vaccine delivery including the DCCT, DCCA and driver who were given as daily allowance. The total monthly costs expended for vaccine delivery were $ 445.9 (Table 8). When we considered the total number of children under one-year in Gomba district (7,290), the $ 445.9 total cost for vaccine delivery under the IPM model translates into a unit cost of $0.06 to reach each infant with vaccination services.

**Table 8:** Monthly costs incurred by district to deliver in vaccines during implementation.

| Cost item | Unit cost in USD | Quantity | Frequency | Total in USD |
| --- | --- | --- | --- | --- |
| Safari Day allowance | 5.4 | 3 | 5 | 81.1 |
| Fuel in liters | 5.4 | 40 | 5 | 270.3 |
| Communication | 5.4 | 1 | 1 | 13.5 |
| Car maintenance | 5.4 | 1 | 1 | 81.1 |
| **Total** |  |  |  | **445.9** |

A total of 5 days was used to deliver vaccines to all health facilities following a pre-defined route. On average 1200 kilometres were travelled within the 5 days of the months’ delivery and approximately 200 litres of fuel were consumed. Therefore, 6 litres of fuel were required for each kilometre moved in distance. The costs budgeted and incurred included safari day allowance, fuel, communication, and car maintenance costs.

As noted, a few financial challenges were highlighted prior to IPM as follows:

> “The main challenge with vaccine delivery is lack of funds and consistence in flow of funding because when you have this time and don’t have in the next then you can’t deliver without funds. The main issue is funds, once you have the funds the rest are easy to work with”. **KII, District Cold Chain Technician, Gomba district.**

## Discussion

This is the first known implementation study in Uganda that has assessed a costed informed push vaccine delivery model with its potential benefits in improving the last mile delivery of vaccines. The results of this study can be used for planning and evidence-based response actions. In our study we considered, several components that can the improve efficiency of the last mile delivery system including timely delivery of vaccines, maintenance of cold chain system, and costing of the last mile logistic system.

Our findings show that the lead time delivery of vaccines from the DVS to all immunizing health facilities is 5 days in each month and yet before, health facilities could take on average two weeks to pick vaccines from the district vaccine store. Evidence from this study suggest that when health facilities picked vaccines, there was no real time temperature monitoring done during transportation and therefore no known visible active cold chain maintenance performed which gravely compromised on the potency of vaccines. However, during the IPM intervention, with a use of a trained cold chain technician, cold chain maintenance was enforced and therefore vaccines delivered in their rightful conditions. In addition, there was reduction on vaccine stock-outs as health facilities received vaccines that they were able to consume after proper quantification as opposed to before when they got vaccines in an ad hoc manner. This study demonstrated varying costs for vaccine pick-up which oftentimes were either out-of-pocket expenses incurred by health workers or costs that were eventually charged to caregivers before receipt of immunization services. Moreso, pick-up of vaccines by health workers would impinge on their time for delivering immunisation services which would lead to missed opportunities for vaccination. The monthly cost of vaccine delivery to all health facilities incurred was $445.9 versus combined average $170.8 incurred by all the 19 health facilities for vaccine pick-up. However, the numerous tangible and intangible benefits when the DCCT delivered vaccines to health facilities indisputably outweighs the total costs incurred for vaccine delivery.

### Vaccine delivery model

Our results showed that 79% and 95% of health workers at baseline and endline respectively preferred push model. This finding confirms that health workers prefer to stay at their respective facilities and execute their mandate of delivering healthcare services at the service delivery point rather than having to routinely travel to collect vaccines from the DVS, also recognizing that majority of the health facilities within this setting are struggling with understaffing and patient overload challenges [22]. This takes the pressure off the health worker involved in picking vaccines from the DVS. The constraints related to low staffing levels at the health facilities is not alien and has been widely documented by several scholars which affects access to vaccination services [23].

These results are similar to study conducted in Nigeria that showed a push vaccine delivery model saved one to six hours of the health care workers’ time spent on vaccine logistics (collection and distribution) each week. This enabled healthcare workers to focus on delivering services to mothers and children with attendant reduction in clients’ waiting time as well as enhanced job satisfaction [10].

Our study showed that 26% (5/19) ‘others’ category that picked vaccines included neighbours and motorcycle riders who represent an untrained, unqualified, and unequipped personnel unfit to handle and manage vaccines during transportation. Most common mode of transport used for transporting vaccines from the DVS to the health facilities were hired motorcycles (for more than half of the facilities). These motorcycles were a more preferred mode of transport because they are flexible, convenience and easily available means of transport regardless of the weather changes, versatile and relatively less costly than hiring or fuelling a car over the same comparative distance to be travelled. In many of the cases, rural areas are disadvantaged due to poor road networks, especially during rainy seasons [24]. However, there are certainly concerns about safety, maintenance of the cold chain for the vaccines, possible theft of vaccines or diversion to other parties. On the other hand, the vaccines could get destroyed in case the motorcycle gets an accident which are quite common in Uganda and are routinely fatal.

Vaccine handling and management is best handled by trained and equipped skilled worker who are able recognise and respond appropriately to temperature excursions [2]. Studies have shown that, poor vaccine handling is often associated with technically incompetent stakeholders [13]. The success of the EPI program is tagged to potent vaccines which depend on good cold chain status, therfore, vaccine management is a critical step [11].

### Lead time

Majority of respondents were not aware of the vaccine delivery schedule by National Medical stores at both baseline and endline. Our results showed that only 5% of the health workers knew the NMS vaccine delivery time schedules and only 11% knew the correct recommended number of days within which vaccines should be delivered to health facilities upon their receipt at the DVS. This is consistent with a study in Ghana which showed that out of the 21 respondents, 6 (29%) had heard of “vaccine supply period,” Of which, 2 (33.3%) knew the purpose of the supply period and could correctly tell the vaccine supply period for their facility store [25].

This suggests that more effort is still needed to bridge the gap the knowledge gap around microplanning and vaccine delivery schedules. It is also important to recognise that health workers at facility level need to have knowledge about the vaccine delivery schedule by national medical stores so that their immunization micro plans synchronize with the vaccine delivery schedules which can be helpful in avoiding vaccine stock-outs [26]. For example, in 2011, the Ministry of Health-Uganda acknowledged that vaccine stock-outs were not due to shortages but rather attributed to persistent distribution problems within the districts [27].

#### Cold chain and vaccine management

The cold chain remains a highly vulnerable point for number of EPI programs in developing countries [28]. The core of vaccine cold chain is temperature monitoring [2]. Since all vaccines need to be maintained within certain temperature ranges, the monitoring of vaccine temperatures becomes a must especially through transportation [2]. All Health workers reported reading VVMS upon receipt of vaccines at the health facility. These results are consistent with results from various vaccine assessments that show that health workers are knowledgeable about VVMs and their use in protection of vaccines from heat excursions [29]. However, VVMs only capture the heat exposure over time and not freezing conditions [2]. Indeed, studies have shown that Cold chain management weaknesses are often observed during transportation and storage of the vaccines [28].

Our study showed that only 6/19 (31.58%) health workers used conditioned icepacks during vaccine pick-up. These results are consistent with a study conducted in Ghana where only 23% of health workers had heard of the “conditioning” ice packs [25].

The DCCT is not only meant to deliver vaccines to the health facility, but also supports the vaccine supply chain system like quality assurance activities related to temperature monitoring to maintain cold chain, cross checking for good functionality of the cold chain equipment at a facility, mentorship of health workers in vaccine management, etc. Storage vaccine equipment was functional in majority of the facilities and they all facilities had functional cold chain equipment at endline. Therefore, maintenance of vaccine cold chain across the facilities was well done, including use of vaccine carriers for storage when go to pick them from the DVS. It is important to recognise that monitoring of the cold chain performance is often limited by lack of performance management systems and are rarely done due to limited prioritization of cold chain assessments like temperature monitoring studies and funding [30]. However, the gaps in cold chain management are still evident in transportation of the vaccines from the DVS to the health facilities, with more than 30% of the respondents noting that their facilities do not use frozen ice packs during transportation of vaccines from DVS to health facilities. Such gaps in cold chain management of vaccines during transportation ought to be effectively addressed and managed to ensure vaccine safety, quality and integrity. Inadequate availability of cold boxes and frozen packs to maintain cold chain for vaccines is disadvantageous to the health sector since it attracts a loss of up to 55% of the medicines due to poor storage management of the vaccines [31].

#### Vaccine stock-outs

Results from our study show that the number of health facilities experiencing vaccine stock-outs reduced from 79% (15/19) at baseline to 37% (7/19) at endline. These results are similar to a study conducted in Nigeria that showed a reduction of vaccine stock-outs from 43% to 0% after streamlining their last mile vaccine supply delivery to informed push model [10].

Vaccine stock-outs often lead to missed opportunities for vaccination, decrease number of children who could have been immunised and create a negative demand for immunization [14]. Missed opportunities for vaccination for children that result from inequitable and poor supply chain systems negatively impact the vaccination coverage and can expose children to morbidity and mortality from vaccine preventable diseases which in the long run negatively impacts the economy of a country [14].

#### Costs

Our study revealed varying baseline costs incurred by health facilities for picking vaccines from the district vaccine store with a monthly total combined cost of $ 170.8 for all the 19 health facilities and a monthly median cost of $ 8.1. However, it is important to note that this cost is underestimated as it doesn’t include opportunity costs such as productivity lost when a health worker leaves their duty station to fetch the vaccines from the DVS. Furthermore, while the monthly median cost was quiet low ($8.1), it is also important to highlight that in a context where resources are highly constrained, most facilities often struggled to mobilize the costs of transportation of vaccines and primarily relied on the grossly insufficient primary health care funds and out-of-pocket charges to patients. Collecting money from patients/caretakers to finance transportation of vaccines among others exposes them to financial ruin as they seek health care services as a result of increased out-of-pocket expenditure to meet medical and non-medical costs.

For the IPM, our study estimated the monthly cost for implementation of the push model at $ 445.9 and this translates to an estimated quarterly and annual cost of $ 1, 337.7 and $ 5,350.8 respectively. The estimated monthly cost per child reached with vaccination services in Gomba district is $ 0.06. This very low per child cost makes financial sense and a strong case for nation-wide scale up of the IPM.

Furthermore, the elevated costs of the push model (more than two and half times that of the combined costs of all health facilities at baseline to pick the vaccines), are countered by the immense benefits associated with the IPM such as timely delivery of vaccines, maintenance of vaccines in their right conditions, mentorships and ad hoc repairs of cold chain equipment and time saved for health workers to attend to immunisation activities.

There are studies that have used third party logistic companies to deliver the vaccines to the last mile often at high costs. A study in Cape Town (South Africa) found that outsourcing vaccine logistics to the private sector reduced delivery and inventory costs, improved adherence to temperature thresholds, reduced delivery delays, improved handling practises, and allowed greater volume flexibility [32].

It is arguable that the use of third company logistics to deliver vaccines to the last mile may be too expensive and not sustainable. Our research shows that a better and sustainable approach would be empowering the district cold chain teams to use the available cars designated for the EPI programs by to distribute vaccines in a timely and equitably manner while offering support supervision in vaccine management, conducting ad hoc cold chain maintenance of vaccine equipment at the health facilities, and ensuring health facilities have the right stocks of vaccines. The DCCT is able mentor health workers in EPI in stock management, vaccine forecasting whilst having visibility on functionality of cold chain equipment at the health facility level.

A robust last mile cold chain and supply chain system will require technical persons who are able to carry out multiple tasks such as repairs of cold chain equipment, mentorships, support supervision, vaccine management in addition to vaccine distribution.

There are some limitations in our study including the fact the patient costs were not included in the scope of this analysis. Moreover, the indirect costs already borne by the government such as health personnel, opportunity costs, and other capital costs were also not included in this analysis. Although the study is informative in terms of the potential to use IPM approach for vaccine delivery, the findings should be treated with caution because it was limited to one district and the cost of IPM may differ from district to district as it is affected by distances and terrains. The sample size of one district also limits this study’s ability to generalize to other settings.

## Conclusion

As per the authors’ knowledge, the study described here is the first of its kind that has attempted to demonstrate an improved and costed last mile immunisation supply chain model that uses district based trained cold chain personnel. To address the challenges of an inefficient and poorly defined last mile vaccine logistics system, our study implemented and costed an informed push model of vaccine delivery from the district to all immunising health facilities. This streamlined vaccine supply chain system ensured vaccine quality, timeliness of vaccine delivery, reduced stock-outs, reduced the burden of HCWs in vaccine pick-up, ensured timely repairs of broken-down cold chain equipment, provided visibility of stock status of health facilities, provided a platform for onsite mentorship and other less tangible spill over benefits. The costings, successes and areas of opportunities described herein can serve as a guide for other last mile vaccine systems in Uganda and other countries with similar contexts. The monthly costs associated with the IPM ($445.9 and $0.06 per child) make financial sense and a strong case for the model and this is further reinforced by the various benefits of last mile delivery. This benefit is critical in hard-to-reach areas that tend to have frequent vaccine stock-outs, missed opportunities for vaccination and generally poor vaccine coverages.

## Data Availability

The data is available and has been uploaded

## Acknowledgement

We wish to extend our appreciation to the institutions and individuals that contributed to the implementation of this research study, that is East Africa Community Regional Centre of Excellence for Vaccine Immunization, and Health Supply Chain Management, University of Rwanda; Makerere University School of Public Health; Health Support Initiatives, Uganda; Ministry of Health-EPI division; Uganda National Medical Stores; Gomba District Health team; in-charges and EPI health workers of Health Facilities in Gomba district; and the field data collection teams.

## Supporting Information

SI Dataset

